# A GWAS of ACE Inhibitor-Induced Angioedema in a South African Population

**DOI:** 10.1101/2024.09.13.24313664

**Authors:** Jacquiline W. Mugo, Cascia Day, Ananyo Choudhury, Maria Deetlefs, Robert Freercks, Sian Geraty, Angelica Panieri, Christian Cotchbos, Melissa Ribeiro, Adelein Engelbrecht, Lisa K. Micklesfield, Michèle Ramsay, Sarah Pedretti, Jonny Peter

**Affiliations:** Division of Allergy and Clinical Immunology, Department of Medicine, Faculty of Health Sciences, University of Cape Town, Anzio Road, Observatory, Cape Town, 7625, Western Cape, South Africa; Allergy and Immunology Unit, University of Cape Town Lung Institute (Pty) Ltd, George Street, Mowbray, Cape Town, 7700, Western Cape, South Africa; Sydney Brenner Institute for Molecular Bioscience, Faculty of Health Sciences, University of the Witwatersrand, 1 Jan Smuts Avenue, Braamfontein, Johannesburg, 2000, South Africa; Faculty of Health Sciences, Department of Medicine, Nelson Mandela University, Gqeberha, South Africa; Western Cape Department of Health, District 6 Day Hospital, 50 Caledon Street, Zonnebloem, Cape Town, Western Cape, South Africa; South African Medical Research Council/Wits Developmental Pathways for Health Research Unit (DPHRU), Department of Paediatrics, Faculty of Health Sciences, University of the Witwatersrand, Johannesburg, South Africa

**Keywords:** Angioedema, Genome-wide association studies, Angiotensin-converting Enzyme Inhibitor

## Abstract

**Background:** Angiotensin-converting enzyme inhibitor-induced angioedema (AE-ACEI) is a life-threatening adverse event and, globally, the commonest cause of emergency presentations with angioedema. Several large genome-wide association studies (GWAS) have found genomic associations with AE-ACEI. However, despite African Americans having a 5-fold increased risk of AE-ACEI, there are no published GWAS from Africa. The aim of this study was to conduct a case-control GWAS of AE-ACEI in a South African population and perform a meta-analysis with an African American and European American population.

**Methods:** The GWAS included 202 South African adults with a history of AE-ACEI and 513 controls without angioedema following angiotensin-converting enzyme inhibitor (ACEI) treatment for at least 2 years. A meta-analysis was conducted with GWAS summary statistics from an African American and European American cohort (from Vanderbilt/Marshfield with 174 cases and 489 controls).

**Results:** No SNPs attained genome-wide significance. However, 26 SNPs in the post-imputation standard GWAS of the South African cohort and 37 SNPs in the meta-analysis were associated to AE-ACEI with suggestive threshold(p-value<5.0×10^−06^). Some of these SNPs were found to be located close to the genes *PRKCQ* and *RIMS1,* previously linked with drug-induced angioedema, and also close to the *CSMD1* gene linked to ACEI cough, providing replication at the gene level, but with novel lead SNPs.

**Conclusions:** Our results highlight the importance of African populations to detect novel variants in replication studies. Further increased sampling across the continent and matched functional work are needed to confirm the importance of genetic variation in understanding the biology of AE-ACEI.

## 1. Introduction

Cardiovascular disease is an exploding epidemic facing low-middle-income countries (LMICs) in Africa, and hypertension is the leading cause of death globally, with the greatest burden of disease in LMICs [1]. ACEI are a class of drugs that inhibit the renin-angiotensin-aldosterone system (RAAS), with a proven reduction in mortality from hypertension, diabetes mellitus, and cardiac failure [2]. They are widely available and affordable, making them critical for use in LMICs [3]. ACEI use is limited by two major adverse events: ACEI-angioedema (AE-ACEI) and cough (ACEI-cough) [4]. AE-ACEI typically involves the face, tongue, or larynx and can be life-threatening in ∼16% of cases [5]. AE-ACEI incidence ranges from 0.2-0.7% in retrospective studies to 6% in prospective clinical trials [4–6]; in the only large multicentre African study (CREOLE), the incidence was 0.7% [7]. AE-ACEI is the most common angioedema presentation to emergency rooms across the world, including South Africa [4, 8]. African Americans have a 5-fold increased risk of AE-ACEI compared to European populations [4], and this has led several international hypertension guideline groups to favour angiotensin receptor blockers (ARBs) over ACEI in African populations. We have recently argued that these recommendations are potentially flawed with dire consequences given the near-complete absence of studies of AE-ACEI across diverse African populations [6]. This work aims to address this important gap.

Several GWAS and candidate gene studies have been conducted for AE-ACEI [9, 10], with the largest meta-analysis of eight of these cohorts recently published and identifying three SNPs at genome-wide significance (*rs*6687813, *rs*35136400, and *rs*6060237 on chromosomes 1, 14, and 21, respectively). Further mapping and gene-based tests suggest that regulatory effects on *the BDKBR2* and *BDKRB1* genes are the most likely underlying mechanisms for the association on chromosome 14. The genes most likely associated with 20 SNPs on chromosome 1 are *F5* and *PROCR*, encoding endothelial protein C receptor [11]. Other candidate genes with reported associations have been linked with the activities of alternative bradykinin metabolising enzymes (*XPNPEP2* (*rs*3788853), *MME* (*rs*989692), immune regulatory pathways (*PRKCQ* (*rs*500766) and *ETV6* (*rs*2724635)). Few of these polymorphisms have been confirmed functionally or replicated across diverse populations. At present, there have been only two candidate gene association studies (with less than 50 AE-ACEI cases and <250 hypertensive patients on ACEI) from Southern Africa that have studied AE-ACEI and ACEI-responsiveness genomics in Sub-Saharan Africa (SSA)[12, 13].

These studies associated SNPs *rs*1042714 in the *ADRB2* gene, *rs*1799722 in the *BDKRB2* gene, and the B₂ receptor -9 allele in the *BDKRB2* gene with AE-ACEI [12, 13]. Furthermore, two lines of evidence support the hypothesis that ACE biology and genomics may vary substantially across the African continent. First, carboxypeptidases, including ACE, are important in shaping the immunopeptidomes of class-I HLA [14], and *Choudhury et al*. [15] found the HLA region to be highly differentiated across African genomic regions. Thus, with epistatic association between HLA and ACE, ACE genomics may vary substantially across SSA populations. Second, polymorphisms in ABO blood group genes have been associated with ACEI-cough, and it is hypothesized that oligosaccharide moieties, acted on by ABO-encoded glycosyltransferases, impact ACE solubility and protease degradation [16]. *ABO* genes have been under substantial selection pressure in Africa due to links with malaria susceptibility [17], and therefore this may be another important mechanism for regional differences in ACE across Africa. This preliminary GWAS from a South African population was aimed at addressing this current research gap.

## 2. Materials and Methods

### 2.1 Ethics Statement

Research was carried out in accordance with the latest update of the Declaration of Helsinki. Written informed consent was obtained from all participants in the AE-ACEI and control cohort. The study protocol was approved by the University of Cape Town Faculty of Health Sciences Human Research Ethics Committee (HREC 057/2020) and the Human Research Ethics Committee (Medical) of the University of Witwatersrand (M2210108).

### 2.2 ACEI-Angioedema Cohort Description

A total of 207 adult patients who had experienced AE-ACEI were recruited retrospectively in the Western (Cape Town area) and Eastern (Mthatha and Gqeberha areas) Cape provinces of South Africa. Patients were defined as having AE-ACEI if they had angioedema while taking an ACEI, with no preceding episodes of angioedema in the absence of ACEI use, and no recurrence of angioedema after removal of the offending ACEI. All cases were reviewed and adjudicated by a clinical expert in allergology. Samples were also obtained from 460 controls who had taken an ACEI for at least two years without signs or symptoms of AE. Participants provided saliva collected in an OG-600 kit (DNA Genotek) that was stored at room temperature until DNA extraction. The data collected included demographics and clinical history. All samples were anonymised and labelled with random study identifiers, and the collected data was de-identified for analysis. Seven cases and 92 controls from the Soweto site (Gauteng province) of the Africa Wits-INDEPTH Partnership for Genomic Research (AWI-Gen) study cohort in the Gauteng province were added to this cohort [18, 19].

### 2.3 Genotyping and Quality Control

Participants from the Western/Eastern Cape cohort provided saliva collected in an OG-600 kit (DNA Genotek) that was stored at room temperature until DNA extraction. DNA was extracted from saliva using prepIT°L2P according to manufacturer instructions (DNA Genotek, cat#PT-L2P). All cases and controls were genotyped on the Infinium^TM^ H3Africa v2 array on an Illumina iScan instrument (Illumina, CA, USA) (https://chipinfo.h3abionet.org) that has a total of 2,225,121 autosomal SNPs. We used GenomeStudio v2, a software provided by Illumina, to assign genotypes to the raw data and evaluate the Infinium assay controls. H3Africa cluster file v2, generated by Illumina, was used to cluster the genotypes. The PLINK [20] plugin provided by Illumina and supported by GenomeStudio was used to convert the genotype data into PLINK data format to allow for further quality controls using PLINK.

A sex discrepancy check was conducted by calculating the X-chromosome homozygosity rate. Any self-identified male that had a homozygosity estimate of F > 0.8 and any self-identified female that had an estimate of F < 0.2 were removed from the analysis. Genotype and individual missingness in the genotype and samples, respectively, were then checked, and all genotypes and samples that had < 0.02 missingness were removed from the dataset. 1,919,455 SNPs and 635 samples were retained after these quality control checks. All our remaining samples were within ±3 standard deviations from the sample heterozygosity rate mean and were all retained after the heterozygosity check. Hardy-Weinberg Equilibrium (HWE) p-values were then calculated from the controls, and all the SNPs with a p-value<1.0×10^-06^ were further removed, retaining 1,914,061 SNPs. The sample relatedness was then checked from an independent set of SNPs that had been pruned for linkage disequilibrium (LD) by calculating the proportion of identity by descent (IBD) relatedness, *pi-hat*, for each pair of samples. The recommended cut-off in GWAS is to use a *pi-hat* threshold of 0.1875 to exclude related samples [21]. However, most of the current GWAS methods are designed to control for sample relatedness, particularly those that implement the mixed linear model’s association (MLMA) [22]. As GCTA-MLMA [23] was to be used for the GWAS analysis, a lenient threshold was set, and one of the pairs of samples with a *pi-hat* > 0.5 was excluded. This retained 616 samples (195 cases and 421 controls). Common SNPs were then extracted from the dataset, and 1,333,573 SNPs that had a minor allele frequency (MAF) > 0.05 were retained. The AWI-Gen data was subjected to the same QC procedure. The Western/Eastern Cape dataset was then merged with the AWI-Gen data. The final analysis includes 944,944 common SNPs, 202 cases, and 513 controls. Samples were then clustered based on both the self-reported race and ethnicity and also by the phenotype status of the participants by principal component analysis (PCA) using GCTA and plotted using GENESIS [24]. **Figures 1A** and **B** show the PCA plots, respectively, while **Table IS** *(supplementary material)* is the demographic table of the study cohort.

**Figure 1:**
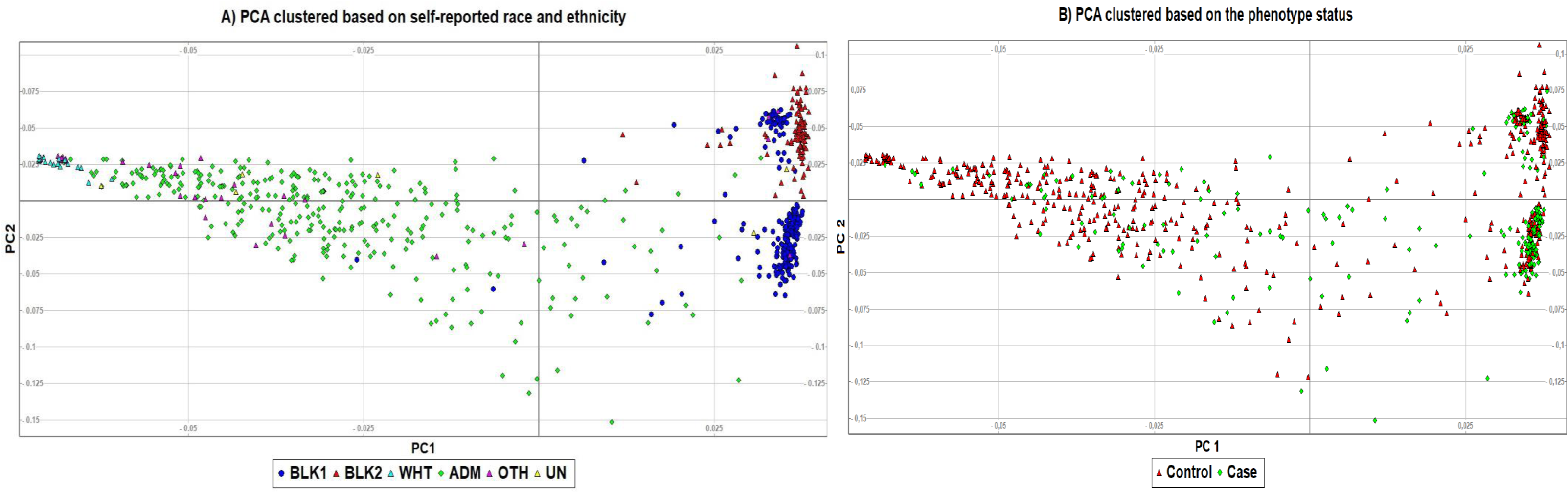
The PCA plot of the South African cohort clustered based on **A)** self-identified race and ethnicity: BLK1 – Black population from Western and Eastern Cape, BLK2 – Black population from Soweto, WHT – White, ADM – mixed ancestry, OTH-Other, UN – Unreported, and **B)** AE-ACEI status. This highlights the genomic diversity and the distribution of the cases and controls in study cohort.

### 2.4 Imputation and GWAS Analysis

The South African cohort was imputed using the African Genome Resource (AGR) panel in the Sanger Imputation Server [25]. This was based on the results of a study of 11,000 sub-Saharan Africans, where >90% of the samples were genotyped using the H3Africa Array, which showed the AGR and Trans-Omics for Precision Medicine (TOPMed) panels performed best in this population [26]. Further quality control was performed on the imputed dataset, where only the SNPs and samples that had <0.02 missingness on the genotype and samples, respectively, and common SNPs with MAF>0.05 were retained. The imputed dataset after quality control had 7,482,056 SNPs and 715 samples. The imputed data was also clustered based on the self-identified ancestry background, and the PCA plot shown in **Figure 1S** (*supplementary material*) was generated. Standard GWAS was then conducted on the imputed dataset using GCTA-MLMA while controlling for age, sex, global ancestry (using 5 principal components (PCs)), and whether or not the samples had hypercholesterolemia, HIV, previous tuberculosis, or asthma, as these were found to be significantly different between the cases and controls (**Table IS**, *supplementary material*). The number of PCs used was determined by obtaining a scree plot for the proportion of variance explained by the first 100 PCs, where 5 PCs were selected using the elbow method (**Figure 2S**, *supplementary material*).

**Figure 2:**
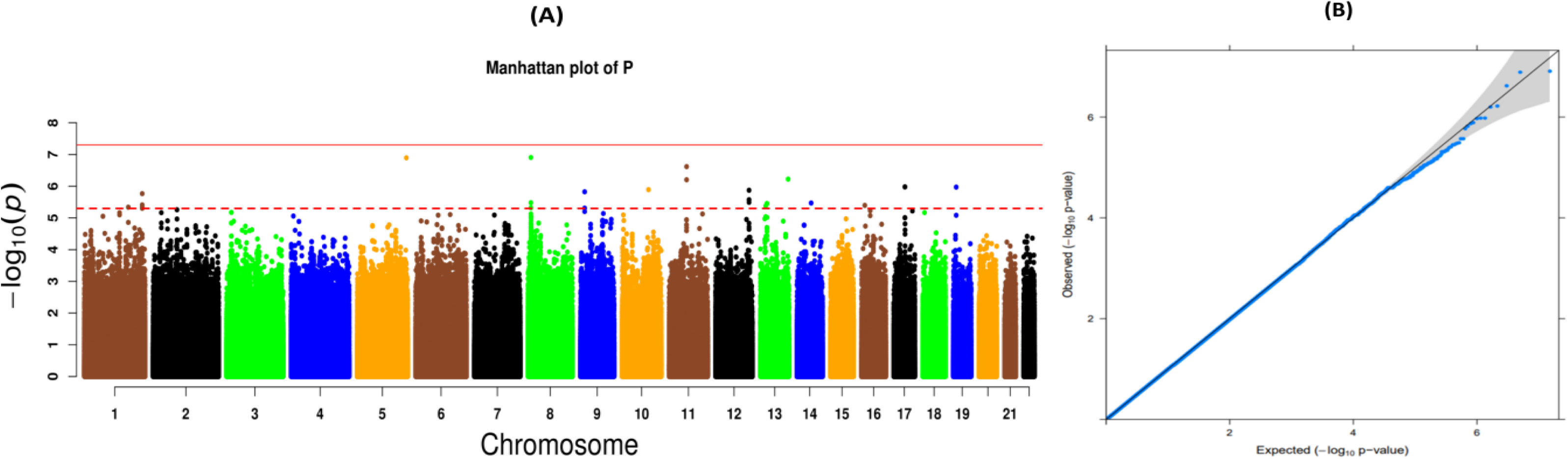
**A)** The Manhattan plot of the standard GWAS of the South African cohort. The dashed red line corresponds to p-value=5.0×10^-06^, while the solid line is the GWAS significance threshold p-value=5.0×10^-08^. **B)** The corresponding QQ plot of the p-values of the standard GWAS (λ=0.99).

### 2.5 Meta-Analysis

A meta-analysis was performed with the South African and American cohort GWAS summary statistics. The data from the Vanderbilt/Marshfield cohort is publicly available by request on dbGAP under study accession number *phs000438.v1. p1*. It consisted of 546,556 autosomal SNPs that had been genotyped using the 610Quadv1.B BeadChip (Illumina, San Diego, California, USA). A build liftover from hg18 to hg19 was first conducted using the liftOver script that is publicly provided by the Centre for Statistical Genetics at the University of Michigan. This cohort was imputed using the Consortium on Asthma among African-ancestry Populations in the Americas (CAAPA) reference panel on the Michigan Imputation Server [27]. The panel consists of 883 African American individuals. After a quality check on the data, where genotypes and samples that had a missingness quality of < 0.02 and rare SNPs with MAF < 0.05 were excluded, 5,222,201 SNPs and the 663 samples were retained. A standard GWAS of this cohort was then conducted using GCTA-MLMA with sex and two PCs as covariates, and genomic control (GC) was performed on the p-values. The total sample size for the meta-analysis was thus 376 AE-ACEI cases and 1002 controls. 4,589,885 SNPs that were in common in the South African and Vanderbilt/Marshfield cohorts were selected. METASOFT [28] was first implemented to estimate the heterogeneity of the study, which was found to be 0, and thus a fixed effect model in METASOFT was used for the meta-analysis.

### 2.6 Functional Annotation and Gene-Based Tests

Functional annotation of the SNPs and prioritization of genes were performed using FUMA (v1.5.2) [29], while MAGMA (Multi-marker Analysis of GenoMic Annotation), implemented in FUMA, was used in the gene-based tests. All the SNPs from the GWAS of the South African cohort and the meta-analysis were annotated. The 1000 genomes Phase3 African population reference panel LD backgrounds were used, and a p-value threshold of 1.0×10^-05^ was set for the lead SNPs in FUMA.

### 2.7 Replication

A number of AE-ACEI association studies on angioedema have been conducted to date [9, 30–33]. In particular, *Liau et al.* [30] have identified more than 10 GWAS for AE-ACEI. Cumulatively, these studies have highlighted a total of 75 SNPs that have been found to be associated with angioedema. We sought to replicate some of the SNPs detected in both the standard GWAS and the meta-analysis.

## 3. Results

The AE-ACEI cases (n=202) and controls (n=513) used in the standard GWAS and meta-analysis were similar in terms of age (55.5±17.8 years vs. 59±18.0 years, respectively, p-value=0.16), predominantly female sex (63% [128/202] vs. 52% [269/513] respectively, p-value=0.008), and co-morbid illness. The AE-ACEI cases had significantly more patients who self-reported as black at 58.6% (116/202), than the ACEI-tolerant controls at 43.3% (224/513) (p-value<0.001) (**Table IS**, *supplementary material*).

### 3.1 GWAS Association and Functional Annotation of the South African Cohort

The Manhattan and QQ plots of the post-imputation South African GWAS are shown in **Figures 2A** and **B**, respectively. In this analysis, 26 SNPs (**Table I**) were detected at a suggestive threshold (p-values<5.0×10^-06^). Additionally, no inflated p-values were observed in the post-imputation GWAS analysis (λ=0.99). As is common in GWAS, most of the 26 SNPs detected at a suggestive threshold were found to be either intergenic or intronic variants and included both imputed and genotyped variants (**Table I**). These were also found to be close to 14 genes by positional mapping. The CUB and Sushi multiple domains 1 gene, *CSMD1*, reported by *Hallberg et al.* [34] in the GWAS catalog, has been linked to ACEI-induced cough. None of the other variants have previously been linked to angioedema.

**Table I:** List of 26 SNPs that were obtained at a suggestive threshold (p-value < 5.0 x 10^-06^) in the post-imputation standard GWAS, the corresponding list of genes they were found close to the SNPs by positional mapping, their function, and whether the SNP was genotyped or imputed.

| CHR | rsID | Position | A1 | A2 | MAF | Pvalue | Nearest Gene | Function | Genotyped/Imputed |
| --- | --- | --- | --- | --- | --- | --- | --- | --- | --- |
| 1 | rs6666273 | 191428753 | A | G | 0.23 | $4.58 \times 10^{-6}$ | *RP11-309H21.2* | intergenic | Imputed |
| 1 | rs4660011 | 241039771 | C | A | 0.44 | $1.72 \times 10^{-6}$ | *RGS7* | intronic | Imputed |
| 1 | rs12239109 | 241042754 | C | T | 0.37 | $4.59 \times 10^{-6}$ | *RGS7* | intronic | Genotyped |
| 1 | rs10926374 | 241047316 | C | T | 0.38 | $4.94 \times 10^{-6}$ | *RGS7* | intronic | Imputed |
| 1 | rs7544945 | 241048562 | T | C | 0.38 | $4.92 \times 10^{-6}$ | *RGS7* | intronic | Imputed |
| 1 | rs12127148 | 241049298 | G | T | 0.37 | $3.9 \times 10^{-6}$ | *RGS7* | intronic | Imputed |
| 5 | rs28630061 | 176204086 | T | C | 0.16 | $1.27 \times 10^{-7}$ | *RP11-375B1.3* | intergenic | Imputed |
| 8 | rs62488207 | 4808991 | T | G | 0.34 | $4.79 \times 10^{-6}$ | *CSMD1* | intronic | Imputed |
| 8 | rs12544185 | 4812064 | A | T | 0.33 | $3.32 \times 10^{-6}$ | *CSMD1* | intronic | Imputed |
| 8 | rs7815832 | 4812436 | A | C | 0.35 | $1.24 \times 10^{-7}$ | *CSMD1* | intronic | Genotyped |
| 9 | rs142454594 | 10278823 | A | G | 0.07 | $1.50 \times 10^{-6}$ | *PTPRD* | intronic | Imputed |
| 9 | rs79552273 | 10279110 | T | C | 0.07 | $4.91 \times 10^{-6}$ | *PTPRD* | intronic | Genotyped |
| 11 | rs12287265 | 60237028 | A | T | 0.20 | $2.41 \times 10^{-7}$ | *MS4A1* | UTR3 | Imputed |
| **11** | *rs75124932* | 60250467 | C | G | 0.20 | 6.28×10<sup>-07</sup> | *MS4A12* | intergenic | Imputed |
| **12** | *rs73409491* | 124545501 | A | G | 0.13 | 3.22×10<sup>-06</sup> | *FAM101A* | intronic | Genotyped |
| **12** | *rs58371925* | 124550518 | C | T | 0.14 | 2.69×10<sup>-06</sup> | *FAM101A* | intronic | Imputed |
| **12** | *rs58449918* | 124551080 | G | T | 0.13 | 2.69×10<sup>-06</sup> | *FAM101A* | intronic | Imputed |
| **12** | *rs112593781* | 124559622 | G | A | 0.14 | 1.34×10<sup>-06</sup> | *FAM101A* | intronic | Imputed |
| **13** | *rs116365021* | 34778472 | G | A | 0.06 | 4.19×10<sup>-06</sup> | *SNORA25* | intergenic | Imputed |
| **13** | *rs28810452* | 38841040 | A | G | 0.19 | 3.53×10<sup>-06</sup> | *UFM1* | intergenic | Imputed |
| **13** | *rs58482084* | 38845507 | G | A | 0.19 | 3.53×10<sup>-06</sup> | *UFM1* | intergenic | Imputed |
| **13** | *rs59296363* | 112101252 | T | G | 0.12 | 6.00×10<sup>-07</sup> | *TEX29* | intergenic | Genotyped |
| **14** | *rs2145156* | 69095279 | A | G | 0.08 | 3.39×10<sup>-06</sup> | *RAD51B* | intronic | Imputed |
| **17** | *rs73321625* | 45977502 | T | A | 0.16 | 1.05×10<sup>-06</sup> | *SP2:AC003665.1* | ncRNA exonic | Imputed |
| **17** | *rs73321628* | 45977632 | G | A | 0.16 | 1.05×10<sup>-06</sup> | *SP2:AC003665.1* | ncRNA exonic | Genotyped |
| **19** | *rs12610494* | 8407470 | C | G | 0.09 | 1.07×10<sup>-06</sup> | *KANK3* | intronic | Imputed |

\***CHR**-chromosome, \***A1**–minor allele, \***A2**-reference alleles, \***MAF**-minor allele frequency

### 3.2 Meta-analysis and Functional Annotation

The Manhattan and QQ plots of the standard GWAS of the Vanderbilt/Marshfield cohort post-imputation are shown in **Figures 3S A** and **B** (*supplementary material*). To correct for the effect of population structure, the p-values were GC-corrected, and the inflation factor improved from λ=0.56 to λ_GC_=1.0. Similar to a previous GWAS on this cohort by *Pare et al.* [35], none of the SNPs studied were observed to be significant; however, 25 SNPs with a p-value<5.0×10^-06^ were detected. Using a pre-imputed dataset of this cohort, *Pare et al.* [35] performed the association study using conditional logistic regression stratified by ancestry and obtained 41 SNPs that had a p-value<1.0×10^-04^. They highlighted two of the SNPs, *rs*500766 on chromosome 10 and *r*s2724635 on chromosome 12, which they linked to the *PRKCQ* and *ETV6* genes, respectively. We observed *rs*500766 at p-value=2.14×10^-05^ and *rs*2724635 at p-value=1.71×10^-03^ in our post-imputation standard GWAS analysis of the cohort. The fixed-effect meta-analysis of the South African and Vanderbilt/Marshfield cohorts resulted in the Manhattan and QQ plots in **Figures 3A** and **B**. No p-value inflation of p-values was observed in the analysis (λ=0.99). The meta-analysis detected 37 SNPs with a p-value<5.0×10^-06^ and were observed to be mainly intergenic or intronic by positional mapping; see **Table II**.

**Figure 3:**
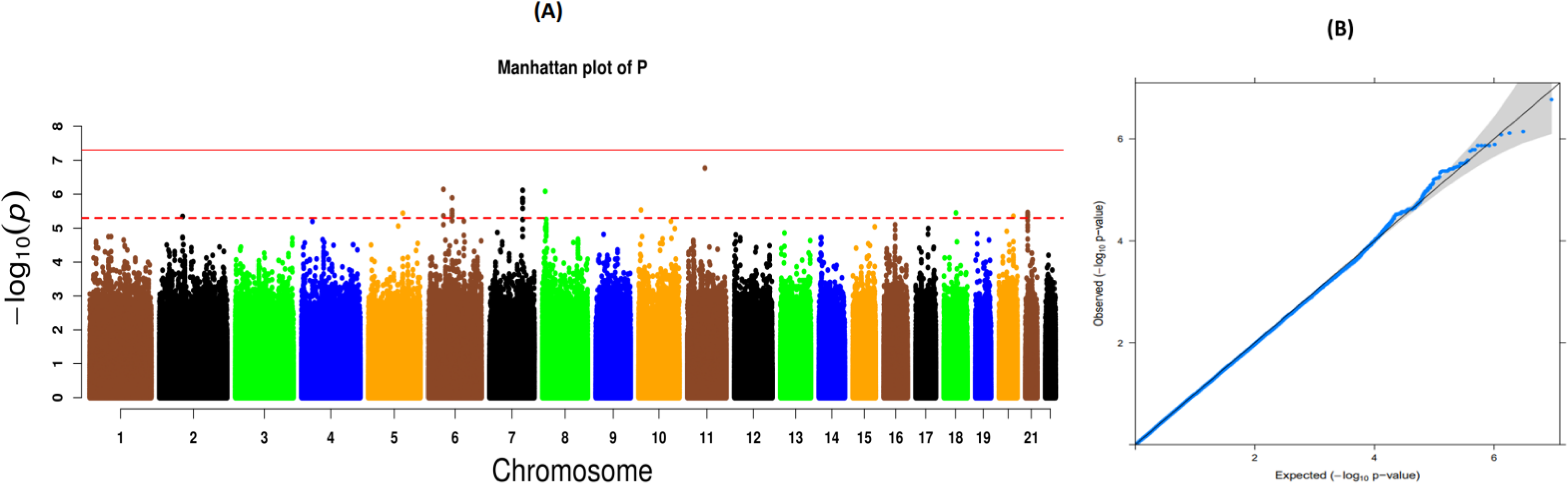
**A)** The Manhattan plot of the meta-analysis of the South African and Vanderbilt/Marshfield cohort summary statistics. The dashed red line corresponds to p-value=5.0×10^-06^, while the solid line is the GWAS significance threshold p-value=5.0×10^-08^. **B)** The corresponding QQ plot of the p-values of the standard GWAS (λ=0.99).

**Table II:** The list of 37 SNPs that were obtained at a suggestive threshold (p-value<5.0 x 10^-06^) in the meta-analysis, the corresponding list of genes they were found close to by positional mapping, their function, and whether the SNP was genotyped or imputed.

| CHR | rsID | Position | A1 | A2 | MAF | P-value | Nearest Gene | Function | Genotyped/Imputed |
| --- | --- | --- | --- | --- | --- | --- | --- | --- | --- |
| 2 | rs6726307 | 72030471 | G | A | 0.2 | $4.51 \times 10^{-06}$ | *DYSF* | intergenic | Genotyped |
| 5 | rs3924097 | 120445067 | T | A | 0.4 | $3.6 \times 10^{-06}$ | *CTD-261308.1* | intergenic | Imputed |
| 6 | rs34388291 | 40580161 | A | G | 0.2 | $4.31 \times 10^{-06}$ | *LRFN2* | intergenic | Imputed |
| 6 | rs34207487 | 40582609 | T | C | 0.21 | $7.23 \times 10^{-07}$ | *LRFN2* | intergenic | Imputed |
| 6 | rs1192177 | 72572886 | T | C | 0.5 | $3.01 \times 10^{-06}$ | ***RIMS1*** | intergenic | Imputed |
| 6 | rs1192178 | 72573194 | A | G | 0.5 | $3.01 \times 10^{-06}$ | ***RIMS1*** | intergenic | Imputed |
| 6 | rs1192179 | 72573244 | C | T | 0.5 | $3.01 \times 10^{-06}$ | ***RIMS1*** | intergenic | Imputed |
| 6 | rs1763305 | 72576793 | G | T | 0.5 | $4.28 \times 10^{-06}$ | ***RIMS1*** | intergenic | Imputed |
| 6 | rs1612461 | 72576794 | A | C | 0.5 | $4.28 \times 10^{-06}$ | ***RIMS1*** | intergenic | Imputed |
| 6 | rs1147527 | 72579416 | A | G | 0.49 | $3.59 \times 10^{-06}$ | ***RIMS1*** | intergenic | Imputed |
| 6 | rs1147526 | 72579596 | G | C | 0.5 | $4.28 \times 10^{-06}$ | ***RIMS1*** | intergenic | Imputed |
| 6 | rs1192182 | 72581129 | G | A | 0.5 | $4.28 \times 10^{-06}$ | ***RIMS1*** | intergenic | Imputed |
| 6 | rs1147522 | 72583600 | T | C | 0.47 | $1.28 \times 10^{-06}$ | ***RIMS1*** | intergenic | Genotyped |
| 7 | rs6952303 | 119603778 | T | C | 0.46 | $1.35 \times 10^{-06}$ | *U1* | intergenic | Imputed |
| **7** | *rs6953016* | 119604064 | A | G | 0.46 | $1.35 \times 10^{-06}$ | *U1* | intergenic | Imputed |
| **7** | *rs10281213* | 119605536 | T | C | 0.46 | $1.74 \times 10^{-06}$ | *U1* | intergenic | Genotyped |
| **7** | *rs7802501* | 119629586 | G | A | 0.46 | $1.35 \times 10^{-06}$ | *U1* | intergenic | Genotyped |
| **7** | *rs10236345* | 119631602 | C | A | 0.46 | $1.35 \times 10^{-06}$ | *U1* | intergenic | Imputed |
| **7** | *rs10257336* | 119635285 | A | G | 0.46 | $1.62 \times 10^{-06}$ | *U1* | intergenic | Imputed |
| **7** | *rs10247779* | 119639646 | G | A | 0.46 | $1.62 \times 10^{-06}$ | *U1* | intergenic | Imputed |
| **7** | *rs58094128* | 119651727 | C | T | 0.46 | $2.62 \times 10^{-06}$ | *U1* | intergenic | Imputed |
| **7** | *rs10245443* | 119656806 | T | G | 0.47 | $7.70 \times 10^{-07}$ | *U1* | intergenic | Imputed |
| **8** | *rs7815832* | 4812436 | C | A | 0.35 | $8.26 \times 10^{-07}$ | ***CSMD1*** | intronic | Imputed |
| **10** | *rs4750617* | 6606334 | A | C | 0.36 | $2.92 \times 10^{-06}$ | ***PRKCQ*** | intronic | Imputed |
| **11** | *rs12287265* | 60237028 | A | T | 0.2 | $1.70 \times 10^{-07}$ | *MS4A1* | UTR3 | Imputed |
| **18** | *rs4553714* | 44298996 | A | G | 0.16 | $3.54 \times 10^{-06}$ | *ST8SIA5* | intronic | Imputed |
| **20** | *rs6021761* | 50776522 | C | A | 0.34 | $4.41 \times 10^{-06}$ | *ZFP64* | intronic | Imputed |
| **21** | *rs2826127* | 21666445 | G | A | 0.38 | $3.45 \times 10^{-06}$ | *AP001171.1* | intergenic | Imputed |
| **21** | *rs9981281* | 21666624 | T | C | 0.38 | $3.45 \times 10^{-06}$ | *AP001171.1* | intergenic | Imputed |
| **21** | *rs1077650* | 21666756 | A | G | 0.38 | $4.27 \times 10^{-06}$ | *AP001171.1* | intergenic | Imputed |
| **21** | *rs1077651* | 21666997 | C | T | 0.38 | $3.89 \times 10^{-06}$ | *AP001171.1* | intergenic | Genotyped |
| 21 | rs1027063 | 21667752 | G | C | 0.38 | $3.89 \times 10^{-06}$ | AP001171.1 | intergenic | Imputed |
| 21 | rs1027064 | 21667841 | T | A | 0.38 | $3.89 \times 10^{-06}$ | AP001171.1 | intergenic | Imputed |
| 21 | rs1027065 | 21667851 | C | T | 0.38 | $4.62 \times 10^{-06}$ | AP001171.1 | intergenic | Imputed |
| 21 | rs1027066 | 21667880 | A | G | 0.38 | $3.89 \times 10^{-06}$ | AP001171.1 | intergenic | Imputed |
| 21 | rs1028981 | 21668483 | A | G | 0.38 | $4.27 \times 10^{-06}$ | AP001171.1 | intergenic | Imputed |
| 21 | rs1028982 | 21668534 | A | G | 0.38 | $4.09 \times 10^{-06}$ | AP001171.1 | intergenic | Imputed |

\***CHR**-chromosome, \***A1**—minor allele, \***A2**-reference alleles, \***MAF**-minor allele frequency.

In addition to detecting SNP *rs*7815832 (p-value=8.26×10^-07^) located near gene *CSMD1* as was observed in the standard GWAS, the meta-analysis detected SNP *rs*4750617 (p-value=2.92×10^-06^) about 56 kb upstream of SNP *rs*500766 located in the *PRKCQ* (protein kinase C theta) gene on chromosome 10 that was linked to AE-ACEI by *Pare et al* [35]. The meta-analysis also detected 9 SNPs at a suggestive threshold on chromosome 6 that were located close to *RIMS1*, the regulating synaptic membrane exocytosis 1 gene, which has previously been linked to angioedema in a Spanish population [36].

### 3.3 Replication Analysis

In this analysis, a Bonferroni-corrected significance threshold of 6.67×10^-04^ (0.05/75) was used. Though SNP *rs*34485356 previously mapped to *BDKRB2* was observed in the standard GWAS with a p-value of 0.004 and SNP *rs*500766 located in gene *PRKCQ* was observed at a p-value of 0.009 in the meta-analysis, none of the SNPs in both studies were replicated at a significant threshold.

### 3.4 Gene-based Test

In total, all the input SNPs in FUMA for the standard GWAS and meta-analysis were mapped to 18,853 and 18,093 protein-coding genes, respectively. The gene-based test using MAGMA for the standard GWAS thus considered a Bonferroni-corrected significance threshold of 2.65×10^-06^ and 2.76×10^-06^ for the two tests, respectively. None of the genes considered were found to be significant. **Tables IIS** and **IIIS** (*supplementary material*) list the top 10 genes highlighted by MAGMA in each analysis, respectively, while **Figures 4S** and **5S** (*supplementary material*) are the corresponding Manhattan plots.

## 4. Discussion

Our results highlight the importance of African populations to detect novel variants and potentially replicate preliminary signals from other populations. To our knowledge, this is the largest GWAS to investigate AE-ACEI in a diverse African cohort on the continent. The underrepresentation of continental African populations in GWAS is concerning [37]. This may lead to healthcare disparities once GWAS results are translated into clinical relevance, as well as limit our understanding of the still-missing heritability that continues to plague GWAS and impact the accuracy of predicting drug responses in diverse populations [38]. In the context of ACEI use, we have highlighted the influence that early epidemiological evidence from African American studies has had on international hypertension guidelines and have warned against the potential pitfalls of extrapolating this very limited data to all continental African populations [6].

In this study, we have performed both a standard GWAS analysis in the South African population and meta-analysed our study with an African American and European American cohort from Vanderbilt (Nashville, Tennessee) and Marshfield (Wisconsin). We have further implemented FUMA to annotate all the SNPs and conducted a gene-based test for the protein-coding genes that were found close to these SNPs. Our standard GWAS of the South African cohort and the meta-analysis detected 26 SNPs and 37 SNPs at suggestive thresholds (p-value<5.0×10^-06^), respectively, which were located close to 23 genes by positional mapping in FUMA. Among them, the *RIMS1* gene has been associated with non-steroidal anti-inflammatory drug (NSAID)-induced angioedema [36]. *RIMS1* modulates G-proteins, in particular those linked to the opening of calcium channels, and this has been best studied in relation to the release of neurotransmitters and insulin [39]. *BDKRB1* and *BDKRB2* are both G-protein coupled receptors, and therefore *RIMS1* may play a role in modulating bradykinin receptor-2 sensitivity in susceptible individuals [40]. In addition, in a GWAS study of smoking patterns and meta-analysis of smoking status, *Xu et al.* [41] found a significant variant *rs*1334346 (p-value=8.22×10^−09^) close to *RIMS1* that was associated with smoking behaviour over time. The *CSMD1* gene, which is a complement regulatory protein linked to kallikrein in pathway analyses, was also found close to the SNPs detected at a suggestive threshold and has been reported by *Hallberg et al.* [34] and *Saunders et al.* [42] on the GWAS catalog.

*CSMD1* is linked to cough in response to ACEI drugs and to age at initiation of smoking, respectively. Smoking has been epidemiologically identified as a risk factor for AE-ACEI [50], but we did not capture smoking status in our clinical data. The *PRKCQ* gene on chromosome 10, which is associated with T-cell activation [43], was a signal highlighted in the original Vanderbilt/Marshfield cohort analysis [35] to be linked to AE-ACEI. As noted in the recent meta-analysis of AE-ACEI in European participants, there is now an urgent need for functional data to confirm the biological role of some of these associated genes, particularly those impacting bradykinin receptor sensitivity and signalling channels.

The main limitation of our study is the small sample size and, thus, the low power to attain genome-wide significance for novel SNP associations. However, these findings highlight the importance of the inclusion of African GWAS in replication and meta-analysis. The large number of SNPs obtained at a suggestive threshold in both the post-imputation standard GWAS and the meta-analysis further highlights the need for increased sampling on the continent if African GWAS is to catch up with European GWAS.

In conclusion, this study presents the largest GWAS of AE-ACEI from a continental African population, with several SNPs detected at a suggestive threshold in both the post-imputation GWAS and meta-analysis, including SNPs near genes with biological plausibility and prior associations with drug-induced angioedema. Further work is now required to increase sampling across diverse African regions to improve study power and further illuminate the heritability of AE-ACEI.

### Perspectives

We performed a GWAS of a South African cohort and a meta-analysis with summary statistics of an African American and European American cohort. We located SNPs associated with AE-ACEI at a suggestive threshold (p-value<5.0×10^-06^) close to the *CSMD1* gene, previously linked to ACEI cough, as well as the *RIMS1* and *PRKCQ* genes linked to drug-induced angioedema. The study highlights the importance of African populations in meta-analysis and replication studies and the need for increased sampling on the continent of Africa.

### Novelty and Relevance What is new?

To our knowledge, this is the largest GWAS and meta-analysis study on AE-ACEI in a South African cohort that has replicated genes linked to drug-induced angioedema.

### What is Relevant?

The study identifies SNPs with suggestive significance thresholds that are associated with AE-ACEI. It also highlights the need for increased sampling on the African continent to ensure novel results and thus equitable healthcare when GWAS hits are translated to clinical relevance.

### Clinical/Pathophysiological Implications?

Identification of risk variants associated with AE-ACEI in African populations will better inform hypertension treatment guidelines for this population.

## Data Availability

The data used in this study are available upon request from the corresponding author.

## Non-standard Abbreviations and Acronyms

AE-ACEI: Angiotensin-converting Enzyme Inhibitor-Induced Angioedema
ACE: Angiotensin converting enzyme
ACEI: Angiotensin converting enzyme inhibitors
ARBs: Angiotensin receptor blockers
AGR: African Genome Resource
AWI-Gen: Africa Wits INDEPTH Partnership for Genomic Research
CREOLE: *<u>C</u>*ompa*<u>r</u>*ison of thr*<u>e</u>*e c*<u>o</u>*mbination therapies in *l*ow*<u>e</u>*ring blood pressure in Black Africans
GCTA: Genome-wide complex trait analysis
GWAS: Genome-wide association studies
H3Africa: Human heredity and health in Africa
LMICs: Low-middle income countries
MAGMA: Multimarker analysis of genomic annotation
MLMA: Mixed linear model’s association
QQ: Quantile-quantile
RAAS: Renin angiotensin-aldosterone system (RAAS)
SSA: Sub-Saharan Africa (SSA)
TOPMed: Trans-Omics for Precision Medicine

## Acknowledgements

We are grateful to the National Integrated Cyber Infrastructure System (NICIS) for their Centre for High Performance Computing in South Africa (CHPC) and the University of Cape Town’s High-Performance Computing (HPC) facilities for providing the computing platform for this study.

## Supplementary data

Supplementary Figures (**1S**-**5S**) and Tables (**IS**-**IIIS**) are available.

